# Prediction of acetaminophen-induced hepatotoxicity in acetylcysteine-treated patients using routine admission biomarkers

**DOI:** 10.64898/2026.07.16.26358122

**Authors:** Chris Humphries, Alastair M. Kilpatrick, Melisande L. Addison, Jennifer A. Cartwright, Marcus J. Lyall, Linus J. Schumacher, Stuart J. Forbes, James W. Dear

**Affiliations:** Institute for Neuroscience and Cardiovascular Research, University of Edinburgh, Edinburgh, United Kingdom; Centre for Precision Cell Therapy for the Liver, University of Edinburgh, Edinburgh, United Kingdom; Centre for Regenerative Medicine, University of Edinburgh, Edinburgh, United Kingdom; The Royal (Dick) School of Veterinary Studies, University of Edinburgh, Edinburgh, United Kingdom; Usher Institute for Population Health Sciences, University of Edinburgh, Edinburgh, United Kingdom

**Author notes:** **Corresponding author:** Chris Humphries, Institute for Neuroscience and Cardiovascular Research, University of Edinburgh, Little France Crescent, Edinburgh EH16 4TJ, United Kingdom.

**Keywords:** acetaminophen, paracetamol, hepatotoxicity, acetylcysteine, machine learning, risk prediction, drug-induced liver injury, emergency medicine

## Abstract

**Study objective:** Some acetaminophen-overdose patients develop hepatotoxicity despite acetylcysteine treatment. Established tools struggle to prospectively identify this cohort. We developed a model using only routine admission biomarkers to identify acetylcysteine-treated patients at highest risk, and compared it with the current benchmark, the alanine aminotransferase × acetaminophen product (ALT×APAP).

**Methods:** Retrospective cohort of all acetaminophen overdose admissions (ICD-10 T39.1) to three UK hospitals (2008–2024) with alanine aminotransferase (ALT) ≤1000U/L at admission. We fitted elastic-net logistic models stratified by presentation ALT. The outcome was peak ALT >1,000U/L. Performance was assessed on a 25% held-out test set and benchmarked against ALT×APAP.

**Results:** Of 4,705 admissions, 119 (2.5%) developed hepatotoxicity. The model used seven routine blood tests, four per stratum: acetaminophen, sodium, potassium and lymphocyte count where presentation ALT was ≤50U/L; ALT, bilirubin, alkaline phosphatase and lymphocyte count where it was 51–1,000U/L. In the test set (n=1,175) it achieved an area under the curve of 0.93 (95% CI 0.89–0.97) versus 0.82 (0.72–0.91) for ALT×APAP (paired difference 0.11; 95% CI 0.01–0.22; p=0.03), with higher specificity and a higher positive likelihood ratio at every matched sensitivity. Matched to current ALT×APAP >1,500 practice (sensitivity 89.7%), specificity was 82.5% versus 62.6% and the positive likelihood ratio 5.1 versus 2.4, more than halving false-positive escalations (171 versus 365 per 1,000 patients).

**Conclusion:** A stratified model using only routine admission biomarkers identifies acetylcysteine-treated patients at highest residual hepatotoxicity risk, outperforming the ALT×APAP rule across decision thresholds, supporting selection for intensified therapy.

**Editor’s Capsule Summary:** What is already known. Acetaminophen (paracetamol) overdose is one of the commonest causes of acute liver injury and acute liver failure presenting to emergency departments. Acetylcysteine prevents hepatotoxicity in most treated patients, but a minority still progress despite treatment. Guidelines describe intensified options for these highest-risk patients (increased-dose acetylcysteine, adjunctive fomepizole, earlier transplant referral) but note that supporting data are limited, and current tools struggle to identify, at presentation, which acetylcysteine-treated patients will progress.

What this study adds. A model using only seven routine admission blood tests identified those at highest residual risk of hepatotoxicity (peak ALT >1,000U/L) with an area under the curve of 0.93 on a held-out test set. It outperformed the current two-variable rule across the range of thresholds examined, providing higher specificity and a higher positive likelihood ratio for any given sensitivity; matched to current practice it more than halved false-positive escalations.

How it is relevant. Because every input is measured on the first routine blood draw after an acetaminophen overdose, the score can be computed at the point of the initial treatment decision and read against whichever risk threshold a given intervention warrants, helping identify which acetylcysteine-treated patients merit intensified therapy or closer monitoring.

## Introduction

### Background

Acetaminophen (paracetamol) overdose is a common reason for presentation to emergency departments following self-poisoning and is a leading cause of acute liver injury, acute liver failure, and emergency liver transplantation.[1–4] Acetylcysteine, given according to weight- and timing-based protocols, is a highly effective antidote that prevents hepatotoxicity (peak alanine aminotransferase greater than 1,000U/L; ALT) in the large majority of treated patients.[5,6] A clinically important minority nonetheless progress to hepatotoxicity, and it is from this hepatotoxic group that the most serious consequences of acetaminophen overdose (acute liver failure, transplantation and death) overwhelmingly arise.[7] These are the patients in whom earlier intervention could conceivably change outcomes. Currently, at the time acetylcysteine treatment begins, clinicians have limited options for identifying which patients will subsequently develop significant liver injury.

### Importance

International guidelines describe intensified options for patients at highest residual risk, such as increased-dose acetylcysteine, adjunctive fomepizole, and earlier referral for transplant assessment, but do not endorse any of them unreservedly and offer no validated way to identify those patients prospectively. The US and Canada consensus statement explicitly notes that “practice varies and few data are available to guide management.”[6] Currently, the best-established tool is the multiplication product of presentation ALT and acetaminophen concentration (the ALT×APAP product), which has externally validated thresholds.[8,9] However, in routine practice the rule flags a large proportion of patients who will not develop hepatotoxicity.[10]

Individual novel biomarkers have been proposed but are unlikely, on their own, to sufficiently resolve this uncertainty, and most are not currently measured as part of routine care.[11–15] While the performance of a two-variable rule can be established statistically, there is also a pathophysiological limit. Presentation ALT and acetaminophen concentration capture the toxic exposure and evidence of established hepatic injury, but no other patient factors that may be associated with injury progression. Several of the tests already routinely obtained at presentation may offer additional predictive value: the full blood count reflects the systemic inflammatory and immune response; routine biochemistry may demonstrate early renal involvement and electrolyte disturbance which may accompany serious poisoning; and the wider liver and coagulation panel reflects hepatic synthetic and excretory capacity.[7,16-18]

A pragmatic alternative is therefore to extract any additional signal from the blood tests that are already drawn at presentation into a single multivariable estimate of residual risk, adding no new assays and requiring no information beyond what is available when acetylcysteine is started.

### Goals of this investigation

We aimed to develop and internally validate a multivariable model that operates after the decision to start acetylcysteine, uses only routine admission blood tests, and identifies acetylcysteine-treated patients at highest residual risk of hepatotoxicity. The objective was to extract any previously unrecognised value from current routine blood tests in order to support selection for intensified therapy or closer observation, with benchmarking against the current best objective comparator (ALT×APAP).

## Materials and Methods

### Design and setting

Retrospective cohort study of all adult and pediatric patients admitted for acetaminophen overdose to three National Health Service hospitals in NHS Lothian (UK) between March 2008 and October 2024. NHS Lothian is a health board region serving a population of approximately 1,000,000 people.

### Selection of participants

Admissions were identified at data extraction by a primary or secondary ICD-10 code of T39.1 (poisoning: 4-aminophenol derivatives). We then distinguished two cohorts. An outcome-validation cohort (all T39.1 admissions) was used only to confirm that the hepatotoxicity endpoint is clinically meaningful. The modelling cohort comprised admissions whose first ALT was ≤1,000U/L and the first ALT obtained in the 24 hours before admission to the inpatient ward (i.e. during the emergency department stay). Admissions already meeting the outcome at the first ALT were excluded from modelling.

Blood tests obtained after admission to the ward were only used to identify outcome information and characterise patient cohorts, because they are not part of the emergency department presentation. Where a patient had several admissions, we retained one randomly, prioritising admissions with severe liver injury. The cohort derivation and the validation of the hepatotoxicity outcome are detailed in Appendix E1.

### Measurements

Candidate predictors were standard admission laboratory values available within ±2 hours of the first ALT in >75% of patients: acetaminophen concentration, ALT, alkaline phosphatase, basophils, bilirubin, creatinine, eosinophils, hemoglobin, international normalized ratio (INR), lymphocytes, monocytes, neutrophils, potassium, sodium and urea, plus age and sex. These tests reflect standard paracetamol overdose admission panels.[16] We used only the blood results available when the first treatment decision was made.

The candidate set reflects routine care rather than a curated selection; elastic-net regularisation then resolves the redundancy among correlated biomarkers. Missing values were handled by multiple imputation by chained equations (50 imputations), fitted on the training set only and re-fitted within each cross-validation fold.[19] The full statistical methods are set out in Appendix E2.

### Outcomes

The primary outcome was hepatotoxicity (peak ALT >1,000U/L) occurring after the first ALT. We validated this endpoint by its discrimination of acute liver failure and all-cause mortality in the outcome-validation cohort.

### Analysis

We used elastic-net logistic regression, which automatically selects the most informative predictors as part of model fitting while preserving an interpretable equation.[20] Patients were split 75%/25% into training/test sets, stratified on the outcome; the test set was held out from all development. We stratified by presentation ALT at the local upper limit of normal (50U/L) into pre-injury (ALT ≤50U/L) and early-injury (ALT 51–1,000) groups. This two-stratum structure was derived by recursive partitioning, a classification-tree method, which identified presentation ALT as the principal split between patients with no biochemical injury at presentation and those with established early injury.[21] Models were fitted across all 50 imputed datasets combined, with class-balanced weighting and 10-fold cross-validation grouped by patient; outputs were calibrated within each stratum by Platt scaling.[22] The full model specification, including the candidate set and the fitted equations, is given in Appendix E3.

To justify the number of predictors selected, we examined nested models of increasing size across five performance domains (discrimination, calibration, overall accuracy, clinical utility, and incremental risk refinement) using patient-level cross-validated predictions. The size was chosen on discrimination alone: we applied the one-standard-error rule to select the smallest model whose cross-validated discrimination lay within one standard error of the best, and then examined how that choice performed across the other domains (Appendix E4).[23,24] To avoid optimism, multiple imputation was re-fitted within each cross-validation fold, and the Platt calibration used in this evaluation was likewise cross-validated. This selection used only the training-set cross-validation and never the held-out test set. It fixed the primary model at four predictors per stratum, seven in total. The full size-by-domain results are reported in Figure 3. The test set was only used to evaluate the candidate model (and in a sensitivity analysis to establish the relative performance of a full 17-biomarker model; Appendix E4).

The comparator was the ALT×APAP product, treated as positive when >1,500, the value at which intensified treatment would typically be started.[8,9] We report threshold-independent comparisons, the area under the receiver operating characteristic curve (AUROC), and model operating characteristics at a range of sensitivities. The two areas under the curve were compared by a paired stratified bootstrap of 2,000 replicates (pROC), because with rare events the normal approximation underlying DeLong is unreliable. We additionally report calibration, decision-curve net benefit, integrated discrimination improvement, and reclassification of patients relative to ALT×APAP, in the Appendices E5–E8.[25,26] The analysis code and the materials needed to reproduce the modelling approach and its evaluation are openly available at Zenodo (doi: 10.5281/zenodo.21358187). A duplicate-inclusive sensitivity analysis (including patient-level clustering for all admissions, rather than just a single admission per-patient), missing data sensitivity analysis and an acetylcysteine-protocol comparison were performed post-hoc (Appendices E9–E11).

## Results

### Characteristics

In the outcome-validation cohort (n=6,619 admissions, including 241 presenting with ALT >1,000U/L who were excluded from modelling), peak ALT discriminated acute liver failure (AUROC 0.98, 95% CI 0.97–0.99; 29 events) and all-cause inpatient mortality (0.91, 0.85–0.98; 35 events), supporting the clinical validity of the endpoint (Appendix E1). We identified 4,705 admissions for modelling, of which 119 (2.5%) developed hepatotoxicity (Table 1). Seventeen predictors met the availability threshold and entered the ranking; the selection rule, applied to training data only and separately within each stratum, retained four predictors in each stratum (seven distinct tests).

**Table 1.** Baseline characteristics of the modelling cohort, by outcome.

| Variable | Train: hepatotox (n=90) | Train: none (n=3,440) | Test: hepatotox (n=29) | Test: none (n=1,146) |
| --- | --- | --- | --- | --- |
| Female sex, n (%) | 51 (56.7) | 2210 (64.2) | 19 (65.5) | 742 (64.7) |
| Age, y | 38 (25–51) | 32 (21–46) | 36 (24–44) | 32 (22–45) |
| NAC: SNAP, n (%) | 41 (45.6) | 1493 (43.4) | 14 (48.3) | 501 (43.7) |
| NAC: 21-h, n (%) | 49 (54.4) | 1947 (56.6) | 15 (51.7) | 645 (56.3) |
| ALT, U/L | 128 (46–322) | 19 (14–30) | 95 (53–201) | 20 (14–31) |
| Alk. phos, U/L | 78 (66–97) | 78 (64–96) | 79 (66–103) | 78 (64–97) |
| Basophils, $\times 10^9$/L | 0.02 (0.01–0.04) | 0.03 (0.02–0.04) | 0.02 (0.01–0.03) | 0.03 (0.02–0.05) |
| Bilirubin, $\mu\text{mol/L}$ | 17 (10–25) | 7 (5–10) | 14.5 (7.5–23.5) | 7 (5–10.5) |
| Creatinine, $\mu\text{mol/L}$ | 74 (63–87) | 68 (61–77) | 75 (64–93) | 67 (60–75) |
| Eosinophils, $\times 10^9$/L | 0.03 (0.01–0.07) | 0.09 (0.03–0.17) | 0.02 (0.01–0.06) | 0.09 (0.03–0.18) |
| Haemoglobin, g/L | 146 (138–157) | 140 (130–150) | 141 (131–150) | 140 (130–150) |
| INR | 1.1 (1.1–1.3) | 1.0 (1.0–1.1) | 1.1 (1.1–1.4) | 1.0 (0.9–1.1) |
| Lymphocytes, $\times 10^9$/L | 1.29 (0.92–1.80) | 2.15 (1.56–2.87) | 1.14 (0.80–1.64) | 2.19 (1.59–2.86) |
| Monocytes, $\times 10^9$/L | 0.70 (0.43–1.02) | 0.58 (0.43–0.76) | 0.63 (0.41–0.97) | 0.57 (0.43–0.74) |
| Neutrophils, $\times 10^9$/L | 7.8 (5.8–10.4) | 5.1 (3.7–7.3) | 7.3 (5.7–10.4) | 5.0 (3.6–7.5) |
| Paracetamol, mg/L | 100 (37–196) | 63 (23–121) | 129 (48–282) | 72 (22–131) |
| Potassium, mmol/L | 3.8 (3.4–4.1) | 3.9 (3.6–4.1) | 3.6 (3.4–3.9) | 3.9 (3.6–4.1) |
| Sodium, mmol/L | 138 (135–140) | 140 (138–142) | 138 (136–140) | 140 (138–142) |
| Urea, mmol/L | 4.3 (3.5–6.2) | 3.6 (2.8–4.6) | 4.3 (3.6–5.4) | 3.6 (2.8–4.6) |
| Peak ALT, U/L | 2700 (1891–6484) | 21 (15–36) | 6004 (2300–8786) | 22 (14–37) |

Values are median (IQR) unless stated. The four predictors of the primary model per stratum are acetaminophen, sodium, lymphocytes and potassium (pre-injury) and alkaline phosphatase, ALT, lymphocytes and bilirubin (early injury).

### Number of predictors

In both strata discrimination rose quickly as predictors were added, and then plateaued. The one-standard-error rule (a standard way of selecting the simpler model) selected models built from four predictors in each stratum, with the number of predictors selected independently within each stratum. In the test set, the model reached AUROC of 0.93 (statistically indistinguishable from a model using all 17 biomarkers at 0.94), confirming on held-out data the choice made on training data alone (Figure 3). The primary model therefore uses four tests in each stratum, seven distinct routine blood tests in all: acetaminophen, sodium, lymphocyte count and potassium in the pre-injury stratum, and alkaline phosphatase, ALT, lymphocyte count and bilirubin in the early-injury stratum.

### Discrimination and calibration

In the held-out test set (n=1,175; 29 events) the primary model achieved AUROC 0.93 (95% CI 0.89–0.97) versus 0.82 (0.72–0.91) for ALT×APAP. The two marginal intervals describe how precisely each area is estimated; whether the areas differ is assessed by their paired difference, 0.11 (95% CI 0.01–0.22; paired bootstrap p=0.03). All three quantities come from one paired stratified bootstrap; because both scores are evaluated on the same patients their errors are correlated, so the difference is estimated more tightly than the marginal intervals suggest.

The model’s receiver operating characteristic curve lay above every published ALT×APAP operating point, so for any sensitivity a clinician might choose the model returned it with higher specificity (Figure 2A; Appendix E5). Calibration varied across the two strata and was limited by event rate, so benchmarking of the model against ALT×APAP used sensitivity and specificity.

**Figure 1.**
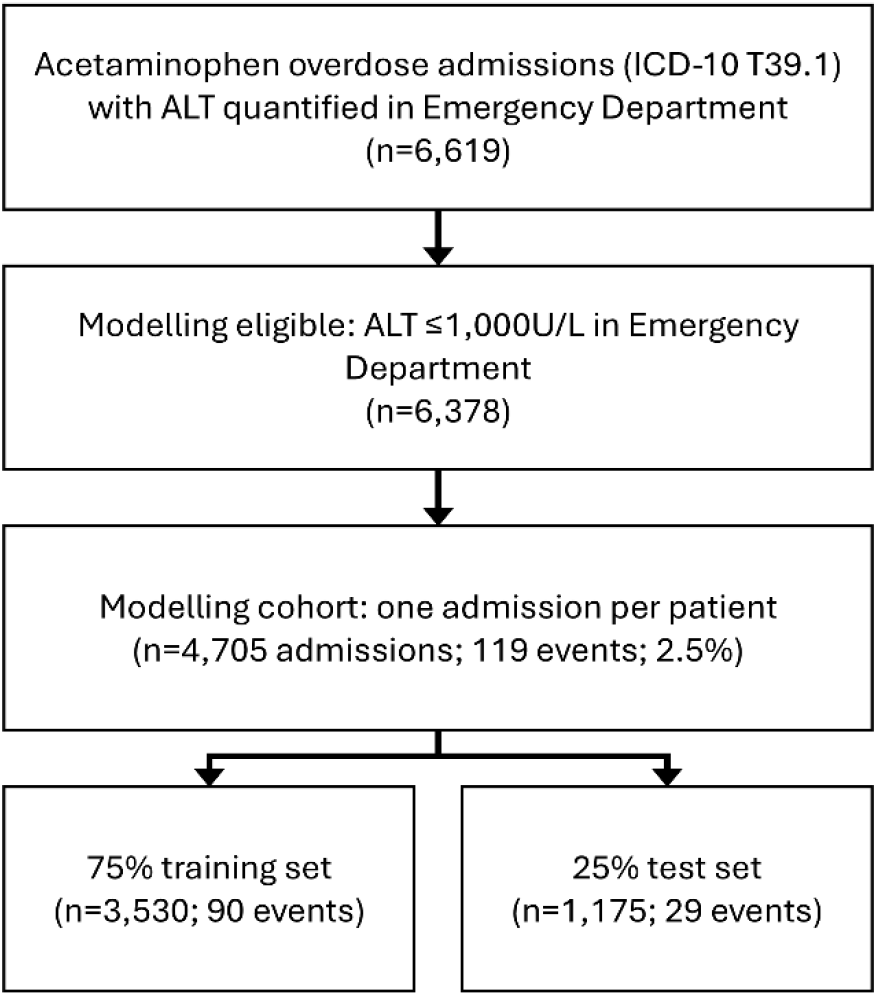
Study flow diagram.

**Figure 2.**
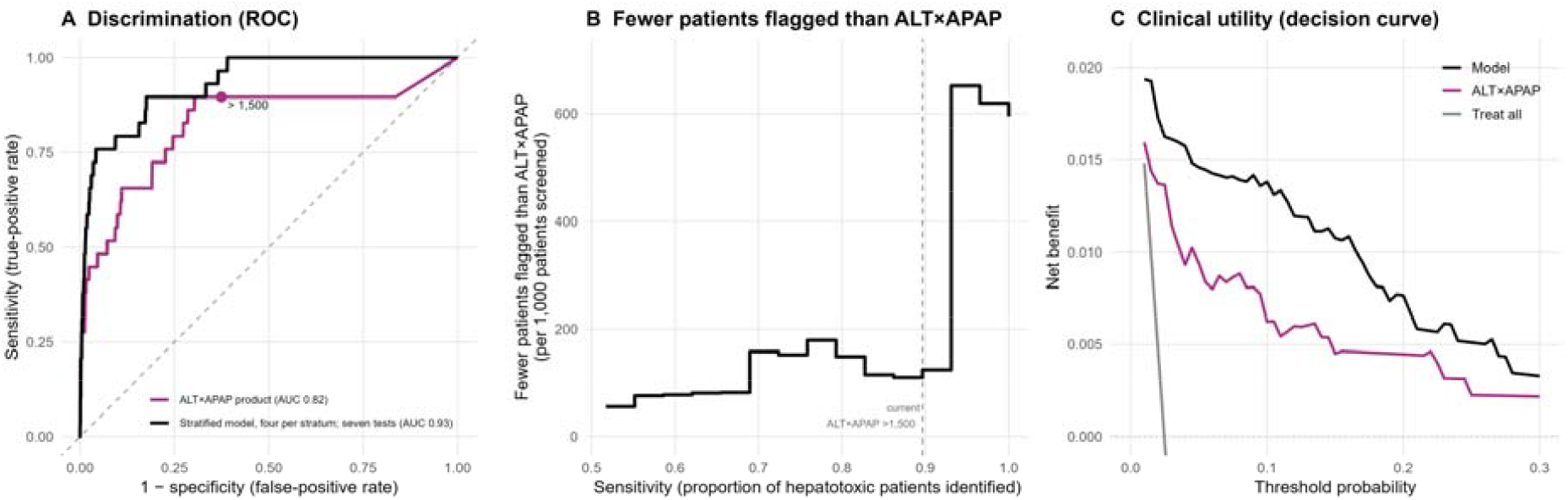
Held-out test-set performance. (A) ROC for the primary model (AUC 0.93) and ALT×APAP (AUC 0.82); the current ALT×APAP >1,500 operating point is marked. (B) Patients spared from being flagged, per 1,000 screened, by using the model instead of ALT×APAP at the same sensitivity — about 194 fewer per 1,000 at current ALT×APAP >1,500 practice, rising at higher sensitivities where ALT×APAP must flag almost every patient. (C) Decision-curve net benefit, shown as supporting evidence given the calibration uncertainty noted in the text.

**Figure 3.**
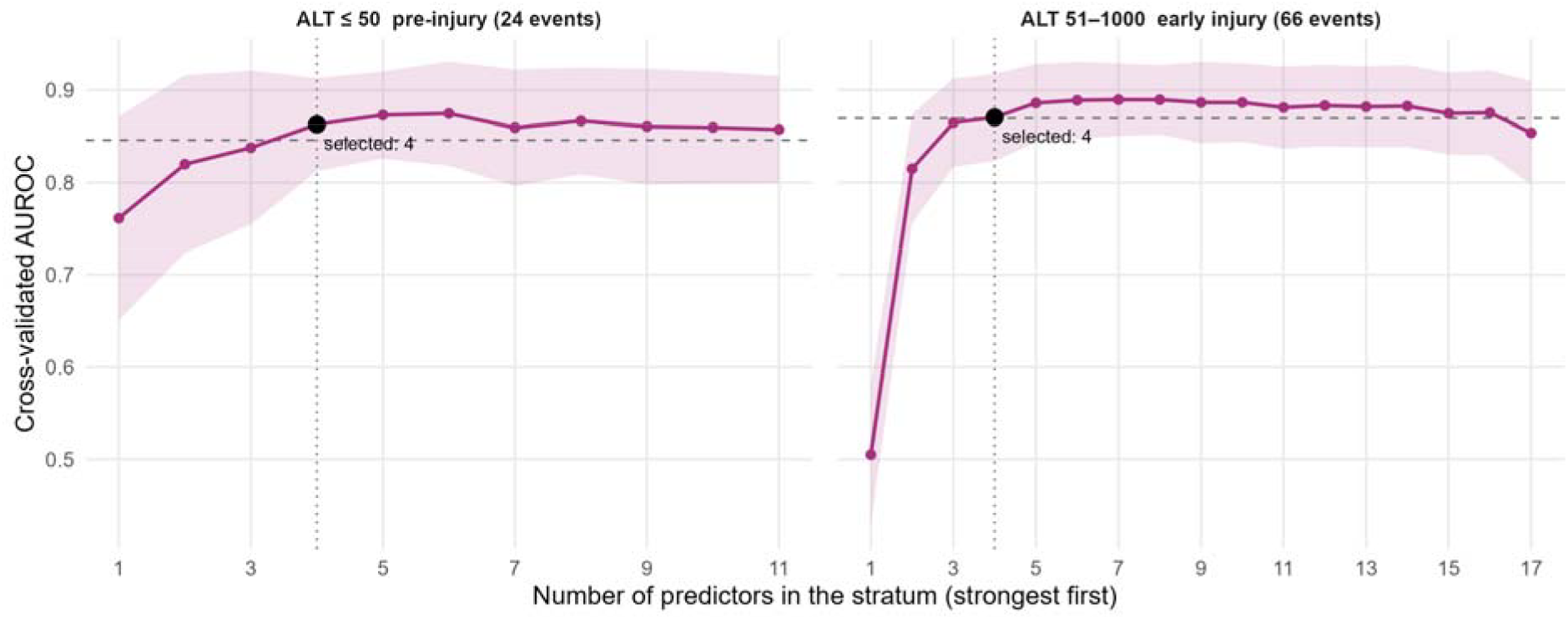
Cross-validated discrimination (AUROC) versus number of predictors, by stratum. The shaded band is the 95% confidence interval; the dashed line marks one standard error below the best size; the filled point marks the size chosen by the one-standard-error rule (four per stratum). Training cross-validation only; the held-out test set was not used in this selection.

### Clinical utility

Across the range of credible decision-threshold probabilities, decision-curve analysis showed the model’s net benefit exceeded that of ALT×APAP (Figure 2C; Appendix E7). Net benefit expresses, in a single common currency, the true-positive gain of a decision strategy after subtracting its false positives weighted by the odds of the chosen threshold; a higher net benefit means that, at that threshold, using the model to decide whom to escalate would identify more patients who go on to progress without a net excess of unnecessary escalations.

### Risk refinement

Against ALT×APAP, the integrated discrimination improvement was 0.13, a threshold-free measure of how much more the model separates the predicted risks of those who progress from those who do not, computed across the whole risk range rather than at any single cut-off; the categorical reclassification of patients relative to the rule, including its event and non-event components, is reported in Appendix E6. We do not lead with the categorical net reclassification index because performance is assessed at an arbitrary risk cut-off and, once the model and the rule are compared at a matched sensitivity, its event component is fixed by construction; the threshold-free measure represents the added discrimination more faithfully.[27]

### Operating characteristics

Because the model returns a continuous risk estimate, a patient’s value can be read against whichever sensitivity a given decision warrants. We compared with the current ALT×APAP >1,500 at matched sensitivity (89.7%, 95% CI 73.6-96.4). At matched sensitivity the model flagged 227 patients to the rule’s 455, more than halving the false-positive escalations (171 versus 365 per 1,000 patients), raising specificity from 62.6% (95% CI 59.7-65.3) to 82.5% (80.2-84.6) and the positive likelihood ratio from 2.39 to 5.11 (Table 3). Because ALT×APAP >1,500 is a single fixed threshold, capturing the patients it misses would require flagging almost the entire cohort, whereas the model reaches any given sensitivity at far lower cost (Figure 2B). A menu of operating-point characteristics is given in Table 3 and Appendix E8.

### Sensitivity and subgroup analyses

A duplicate-inclusive analysis retaining all 6,378 admissions showed substantial between-patient correlation (a logit-scale patient random-intercept variance of 2.34), confirming that repeat admissions are not independent and supporting the decision to retain one admission per patient; predictor directions were consistent, and magnitudes broadly similar, under both a patient random intercept and patient-clustered standard errors (Appendix E9). In the paediatric subgroup (age <18 years; n=112, of whom 2 developed hepatotoxicity, aged 16 and 17), specificity at the operating point was 0.88 (95% CI 0.81–0.93), consistent with the whole test cohort at the same operating point (82.5%; 95% CI 80.2– 84.6); the small number of events precludes a stable estimate of sensitivity and discrimination.

### Limitations

This study represents internal validation within a single health board; external validation in different populations is essential. We did not perform this in our study because our hospitals share a population and laboratories; within-dataset site splitting would have overstated independence.

While the event count is low, the paired bootstrap comparison of the areas under the curve nonetheless reaches p=0.03. However, low event counts also mean calibration cannot be estimated reliably in the pre-injury stratum, prompting threshold selection on sensitivity (which does not rely on calibration). While we have anchored our comparison to an existing model, we would suggest that in external validation and/or clinical use, thought should be given to what operating characteristics are acceptable for the treatment decision being made.

Missing data were not missing at random: laboratory panels are ordered together, so an unordered full blood count removes all its components as a block (and likewise for urea-and-electrolytes and liver function tests). Multiple imputation by chained equations is suited to recovering such structured missingness using the jointly observed panels and outcome, but residual bias cannot be excluded; we therefore report missingness per variable and performance by data completeness (Appendix E10); because the score requires the four routine tests of a patient’s stratum, it is intended for use when those values are available, and we do not claim validated performance where they must be imputed.

Unmeasured factors (ingestion timing, pattern, co-ingestions, time to acetylcysteine, and reasons for blood sampling timings which do not precisely match regimen timings) are not captured in routine structured data and may confound our analysis; elastic net can retain weak or surrogate predictors that should not be interpreted causally. The cohort is predominantly young and female, reflecting UK self-harm epidemiology.

## Discussion

The central finding is that a stratified model using only routine admission biomarkers can identify, among acetylcysteine-treated patients, those at elevated residual risk of hepatotoxicity. Notably, the model improves on the current rule not at one chosen threshold but across the whole range of decision thresholds, so for any sensitivity a clinician might want, the model delivers it with higher specificity. Matched to the sensitivity of current practice it cuts the number of patients escalated by half (227 versus 455) while identifying the same number of patients who will progress. At the stricter thresholds, which would likely be used for the most resource-intensive therapeutic responses, its specificity advantage widens further. This consistent advantage across the thresholds examined is a stronger basis for adoption than superiority at a single cut-off, which can be an artefact of where the cut-off is placed.

A multivariable score raises the fair question of how many variables are warranted over a simple two-variable rule. We addressed this directly by treating model size as a decision to be justified rather than an assumption, and by showing, on training data alone, that discrimination plateaus once a handful of predictors are included. Our resulting model uses seven routine tests, all already drawn at presentation, and the held-out test set confirmed that this parsimonious model performs as well as one using every biomarker. We note that the logic of our model matches the concept underlying ALT×APAP: APAP concentration carries predictive value in early injury, while ALT carries predictive value in evolving injury. The difference is that our model incorporates additional predictive biomarkers to refine that risk estimate.

For clinical use, the question is which threshold to apply. We deliberately do not mandate one. Because the model’s calibration was imprecise and could be estimated reliably in only one stratum, we do not anchor decisions to absolute individual risk values; instead we define each operating point by the sensitivity it delivers, which depends on how the model ranks patients rather than on the exactness of its probabilities. The threshold matched to current ALT×APAP >1,500 practice is a natural default because it ports what is already done, simply doing it more specifically.

The ideal threshold for application is a health-economic question rather than a purely statistical one: it is the point at which the expected benefit of acting equals the expected harm and cost of acting, and that balance differs by intervention. A low-cost, low-risk response such as closer observation or increased-dose acetylcysteine likely justifies a low threshold and high sensitivity, whereas a costly or higher-risk option such as fomepizole, transplant referral, or emerging cell-based therapies, justifies a higher threshold and so a lower sensitivity. The sensitivity at which the tool should operate is therefore a property of the intervention being considered, not a fixed property of the tool, and a single mandated sensitivity would serve the range of possible interventions poorly. The model yields a single risk estimate that can be read against any of these thresholds, and because the escalation options are applied additively when each is independently indicated (for example, a patient may receive both increased-dose acetylcysteine and fomepizole), the same estimate can inform several distinct decisions, each at its own operating point. Formal cost-effectiveness analysis to locate these thresholds is a priority for prospective work.

The model discriminated hepatotoxicity validly in patients treated with either of the two acetylcysteine regimens used in the United Kingdom, with AUROC 0.94 in the 21-hour subgroup and 0.92 in the SNAP subgroup (Appendix E11), showing that the relationship between the admission bloods and the outcome holds across both regimens.

Because the model is a simple logistic equation (Table 2), it can be computed at the bedside with a calculator: no black-box software or specialist bioinformatics infrastructure is required, and the coefficients are disclosed in full. A working spreadsheet calculator, with a short user guide, is provided as a separate appendix to illustrate this. In routine use it would be better delivered by automatic calculation within the electronic health record from blood results that are already returned, so that a risk estimate is available at the first clinical decision without additional work.

**Table 2.** Primary model coefficients (four predictors per stratum; seven distinct tests), raw-value scale, with worked example.

| Predictor | Pre-injury (ALT $\leq 50$) | Early injury (ALT 51–1000) |
| --- | --- | --- |
| Intercept | 37.26694 | 1.10185 |
| Acetaminophen (mg/L) | 0.01033 | – |
| Sodium (mmol/L) | –0.23192 | – |
| Potassium (mmol/L) | –1.35008 | – |
| Lymphocytes ($\times 10^9$/L) | –0.62822 | –0.91820 |
| Alkaline phosphatase (U/L) | – | –0.01485 |
| ALT (U/L) | – | 0.00836 |
| Bilirubin ($\mu\text{mol/L}$) | – | 0.03379 |
| Platt intercept | –4.68139 | –1.72485 |
| Platt slope | 0.57342 | 0.65420 |

Raw score = intercept + $\Sigma$ (coefficient × value). Probability = $1/(1+\exp(-(\text{Platt intercept} + \text{Platt slope} \times \text{raw score})))$ . Worked example, early injury: lymphocytes 1.0, ALT 200, alkaline phosphatase 90, bilirubin 20 gives raw score 1.20, calibrated logit –0.94, probability 28.0%.

**Table 3.** Operating characteristics on the held-out test set (n=1,175; 29 events): current ALT×APAP >1,500 practice, the model matched to that sensitivity, and a menu of model operating points across sensitivities.

| Operating point | Threshold | Sensitivity % (95% CI) | Specificity % (95% CI) | LR+ | LR– | Flagged (n) |
| --- | --- | --- | --- | --- | --- | --- |
| ALT×APAP >1,500 (current practice) | — | 89.7 (73.6–96.4) | 62.6 (59.7–65.3) | 2.39 | 0.17 | 455 |
| Model, matched to ALT×APAP >1,500 | 0.017 | 89.7 (73.6–96.4) | 82.5 (80.2–84.6) | 5.11 | 0.13 | 227 |
| Model at 75% sensitivity | 0.094 | 75.9 (57.9–87.8) | 95.8 (94.5–96.8) | 18.11 | 0.25 | 70 |
| Model at 80% sensitivity | 0.021 | 82.8 (65.5–92.4) | 84.4 (82.2–86.4) | 5.30 | 0.20 | 203 |
| Model at 85% sensitivity | 0.018 | 86.2 (69.4–94.5) | 82.7 (80.4–84.8) | 4.99 | 0.17 | 223 |
| Model at 90% sensitivity | 0.009 | 93.1 (78.0–98.1) | 66.7 (63.9–69.3) | 2.79 | 0.10 | 409 |
| Model at 95% sensitivity | 0.008 | 96.6 (82.8–99.4) | 63.4 (60.6–66.2) | 2.64 | 0.05 | 447 |
| Model at 100% sensitivity | 0.007 | 100 (88.3–100) | 61.0 (58.1–63.8) | 2.56 | 0.00 | 476 |

Sensitivity and specificity are shown with 95% Wilson confidence intervals. LR+, positive likelihood ratio; LR–, negative likelihood ratio; Flagged, number of the 1,175 test patients identified as high risk; Threshold, calibrated-probability cut. Every model row is read from the same continuous risk score.

The next steps are prospective external validation, evaluation in populations with different case mix, and the cost-effectiveness analysis needed to anchor intervention-specific thresholds. We do not claim the model should determine acetylcysteine duration, triage patients away from acetylcysteine, or serve as a general prognostic tool.

In summary, we developed and internally validated a stratified model (the Edinburgh Risk Score) that discriminates acetaminophen hepatotoxicity in acetylcysteine-treated patients using only routine admission biomarkers. This model outperforms the current two-variable rule across the range of thresholds examined, and it can be computed at the first clinical decision to support selection for intensified therapy.

## Supporting information

Reporting checklist

Edinburgh Risk Score Calculator

## Data Availability

NHS Lothian data underlying this study were provided under Caldicott Guardian approval for the specified research aims. The raw data are not publicly available due to patient confidentiality restrictions. Analysis code (stripped of patient data) is available at Zenodo (https://doi.org/10.5281/zenodo.21358187). Requests for access to the data for new uses would require a new application and approval from the Caldicott Guardian.

## Ethics Statement

This study was reviewed and approved by the Edinburgh Medical School Research Ethics Committee (25-EMREC-070), with data extraction and transfer approved by the NHS Lothian Caldicott Guardian (ref. 24167). Patient and public involvement was not sought for this study.

## Author Contributions

C.H. and J.W.D. conceived and designed the study. C.H. performed the data analysis. C.H., A.M.K., M.L.A., J.A.C., M.J.L., L.J.S., S.J.F., and J.W.D. contributed to data acquisition, analysis, and interpretation. C.H. drafted the manuscript. All authors reviewed the manuscript critically for important intellectual content and approved the final version. C.H. takes responsibility for the paper as a whole.

