## Supplemental for "Prediction of acetaminophen-induced hepatotoxicity in acetylcysteine-treated patients using routine admission biomarkers"

### E1. Cohort derivation and outcome validation

The outcome-validation cohort comprised 6,619 eligible admissions. Restricting to admissions with a presentation ALT at or below 1,000 U/L (the modelling-eligible range) left 6,378 admissions. Removing repeat admissions to retain one admission per patient under the submitted segmentation gave the 4,705-admission modelling cohort, which contained 119 hepatotoxicity events (2.5%). The modelling cohort was split into a training set of 3,530 admissions and a held-out test set of 1,175 admissions; the test set contained 29 events.

| Derivation step | Admissions |
| --- | --- |
| Eligible admissions (outcome-validation cohort) | 6,619 |
| Modelling-eligible (presentation ALT at or below 1,000) | 6,378 |
| Modelling cohort (one admission per patient) | 4,705 |
| Hepatotoxicity events in modelling cohort | 119 (2.5%) |
| Training set | 3,530 |
| Test set (29 events) | 1,175 |

Hepatotoxicity was defined as a peak ALT above 1,000 U/L occurring after the first ALT result. To confirm that this biochemical endpoint tracks clinically severe disease, we tested how well peak ALT discriminated harder outcomes in the full eligible cohort. Peak ALT discriminated acute liver failure with an AUROC of 0.98 (95% CI 0.97 to 0.99; 29 events) and all-cause mortality with an AUROC of 0.91 (95% CI 0.85 to 0.98; 35 events). Acute liver failure was defined by ICD-10 codes K72.0 or K72.9, or K71.x with G92 or G93.4; mortality used the discharge status flag.

| Endpoint | Events | AUROC (95% CI) |
| --- | --- | --- |
| Acute liver failure | 29 | 0.98 (0.97 to 0.99) |
| All-cause mortality | 35 | 0.91 (0.85 to 0.98) |

### E2. Statistical methods

Models were fitted by elastic-net logistic regression (glmnet via caret). Hyperparameters were tuned over alpha in [0.1, 0.9] (nine values) and lambda equal to 10^seq(-5, 0, length 30) by 10-fold cross-validation grouped by patient, maximising the AUROC, with class-balanced weighting and predictor centring and scaling. Missing values were imputed by multiple imputation by chained equations with predictive mean matching, using all 50 imputations via a stacked fit (each imputed row weighted 1/50) to yield a single coefficient set. Model outputs were calibrated to the observed prevalence by Platt scaling of cross-validated predictions. For the model-size selection and this calibration, the multiple imputation was re-fitted within each cross-validation fold, so a held-out fold was imputed only from the training folds and never from itself; the elastic-net penalty was held at the value selected on the whole training set, and the final coefficients were then fitted once on the whole training set. The held-out test set was imputed only from the training-set imputation model. Predictors were retained if available in more than 75% of patients; prothrombin time, estimated glomerular filtration rate, white-cell count and total CO2 were excluded; the warfarin INR field was relabelled INR. All selection, calibration and the predictor-count analysis (section E4) used training-set cross-validation only, with no access to the held-out test set.

The model is stratified by presentation ALT into a pre-injury stratum (ALT at or below 50 U/L) and an early-injury stratum (ALT 51 to 1,000 U/L), each with its own coefficients and its own calibration. The number of predictors was chosen independently within each stratum (section E4). The comparator throughout is the ALTxAPAP product (presentation ALT multiplied by presentation acetaminophen concentration), computed as the imputation-averaged product, the mean of ALT x acetaminophen across all 50 imputations, mirroring how the model probability is averaged across imputations. The comparator is binary, positive when the product exceeds its lower published threshold of 1,500. All model-versus-comparator operating-point comparisons in this appendix are made at matched sensitivity.

### E3. Primary model specification

The primary model uses seven distinct routine admission tests in total, four per stratum, with the number of predictors selected independently within each stratum by the leakage-free rule described in section E4. The equations below take raw (untransformed) laboratory values in their reporting units and return a calibrated probability of hepatotoxicity. A patient is flagged high risk in either stratum when the calibrated probability is at or above the operating point chosen for the decision at hand, but the model is used primarily by ranking rather than by a single fixed cut, and section E8 gives an operating-point menu.

#### Pre-injury stratum (presentation ALT at or below 50 U/L)

Raw score = 37.26694 + 0.01033 x acetaminophen - 0.23192 x sodium - 1.35008 x potassium - 0.62822 x lymphocyte count.

Calibrated logit = -4.68139 + 0.57342 x raw score. Probability = 1 / (1 + exp(-logit)).

Model coefficients (four predictors)

| Predictor | Coefficient |
| --- | --- |
| Acetaminophen (mg/L) | 0.01033 |
| Sodium (mmol/L) | -0.23192 |
| Potassium (mmol/L) | -1.35008 |
| Lymphocyte count (x10^9^/L | -0.62822 |

This stratum uses four routine admission tests.

Calibration constants

| Term | Value |
| --- | --- |
| Intercept | 37.26694 |
| Platt intercept | -4.68139 |
| Platt slope | 0.57342 |

#### Early-injury stratum (presentation ALT 51 to 1,000 U/L)

Raw score = 1.10185 - 0.01485 x alkaline phosphatase + 0.00836 x ALT - 0.91820 x lymphocyte count + 0.03379 x bilirubin.

Calibrated logit = -1.72485 + 0.65420 x raw score. Probability = 1 / (1 + exp(-logit)).

Model coefficients (four predictors)

| Predictor | Coefficient |
| --- | --- |
| Alkaline phosphatase (U/L) | -0.01485 |
| ALT (U/L) | 0.00836 |
| Lymphocyte count (x10^9^/L) | -0.91820 |
| Bilirubin (µmol/L) | 0.03379 |

This stratum uses four routine admission tests.

Calibration constants

| Term | Value |
| --- | --- |
| Intercept | 1.10185 |
| Platt intercept | -1.72485 |
| Platt slope | 0.65420 |

#### Worked example (early-injury stratum)

A patient presents with ALT 200 U/L, alkaline phosphatase 90 U/L, lymphocyte count 1.0 x10^9/L and bilirubin 20 micromol/L. Raw score = 1.10185 - 0.01485(90) + 0.00836(200) - 0.91820(1.0) + 0.03379(20) = 1.20. Calibrated logit = -1.72485 + 0.65420(1.20) = -0.94. Probability = 1 / (1 + exp(0.94)) = 28.0%, a high-risk result.

### E4. Predictor-count sensitivity analysis and model selection

To justify the number of predictors rather than take it on trust, we evaluated models with 1 to 17 predictors per stratum, with the size chosen independently within each stratum. Predictors entered each stratum in order of absolute standardised coefficient in the full model. Every model was assessed across five performance domains: cross-validated AUROC (discrimination), calibration slope, net benefit (clinical utility), and the integrated discrimination improvement of the full model over each candidate (IDI of full vs k). All quantities were computed by patient-grouped cross-validation within the training set only, and calibration used a cross-validated Platt fit, so the calibration assessment is not circular and there is no test-set leakage. Both the predictor ranking and the nested-model evaluation used the training-set cross-validation alone; the held-out test set was never consulted during selection, so the choice of model size could not have been informed by the test result. The number of predictors was chosen by a one-standard-error rule applied to training-set cross-validation only, independently within each stratum, selecting the smallest model whose cross-validated discrimination was within one standard error of the best while keeping calibration and net benefit non-inferior; both strata selected four predictors.

The size was chosen separately within each stratum by the one-standard-error rule applied to training cross-validated AUROC, with non-inferiority margins on calibration, incremental refinement and net benefit. Both strata selected four predictors. The binding constraints were discrimination, through the one-standard-error rule, and incremental refinement, through the integrated discrimination improvement of the full model over each candidate, which fell to about zero by four predictors in each stratum; calibration and net benefit were non-inferior at every size and so did not bind. In each stratum the best cross-validated discrimination occurred at a larger model, six predictors in the pre-injury stratum and seven in the early-injury stratum, but a four-predictor model already lay within one standard error of that best and was therefore chosen; allowing the count to differ between strata mattered in principle, because they carry different numbers of events, but here both settled on four. Carried to the untouched test set, the selected four-per-stratum model reached an AUROC of 0.93, statistically indistinguishable from the full 17-predictor model at 0.94, confirming on held-out data the decisions made on training data alone. Figure 3 of the main text plots this size-versus-performance relationship separately for each stratum.

Cross-validated metrics from the training set only. The one-standard-error rule on cross-validated AUROC, with non-inferiority margins on calibration, refinement and net benefit, selected four predictors in each stratum. Held-out test AUROC: selected four-per-stratum 0.93, full 17-predictor 0.94.

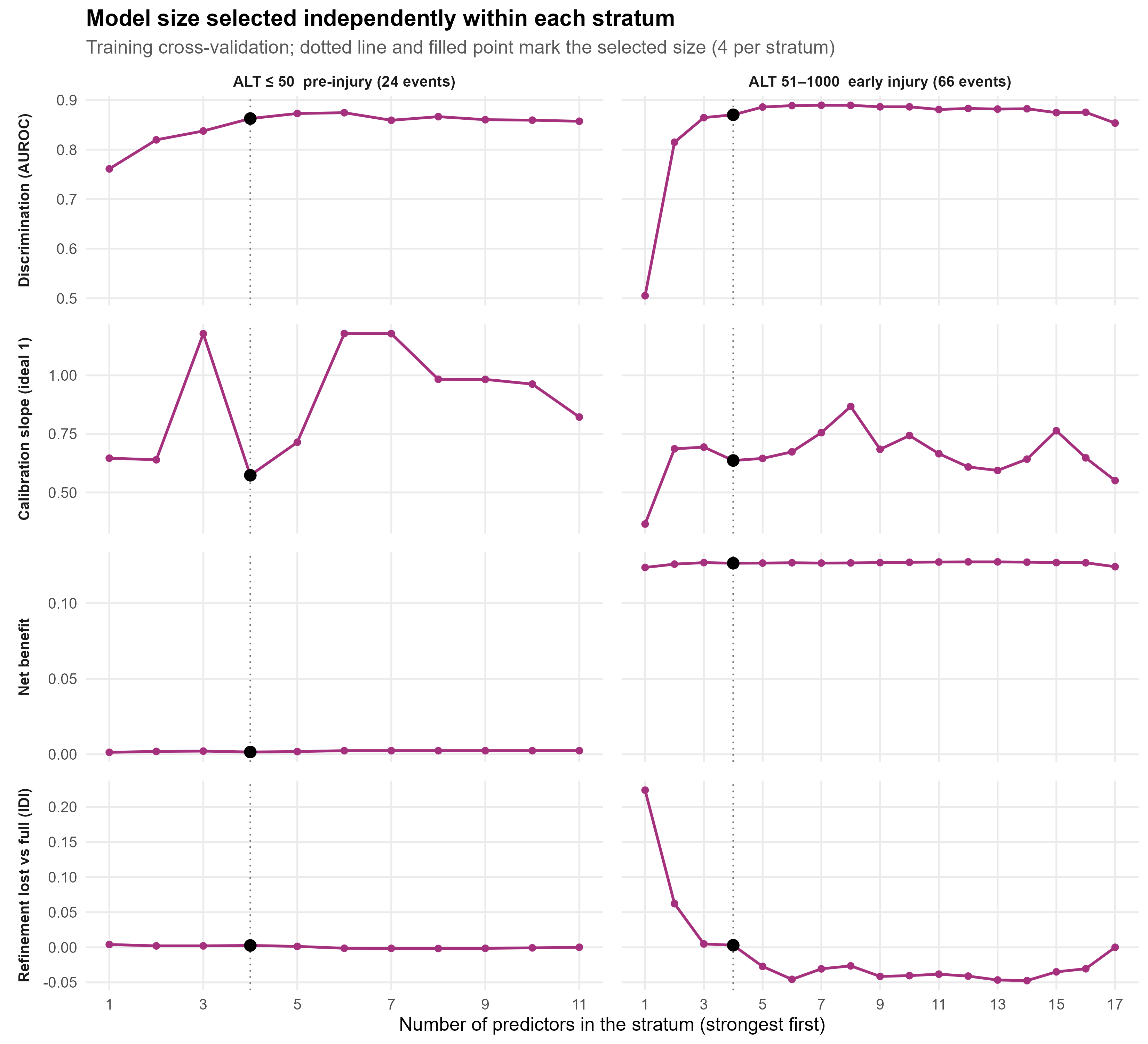

Figure E4. Model-size selection curves by stratum (training cross-validation). Dotted line marks the selected size.

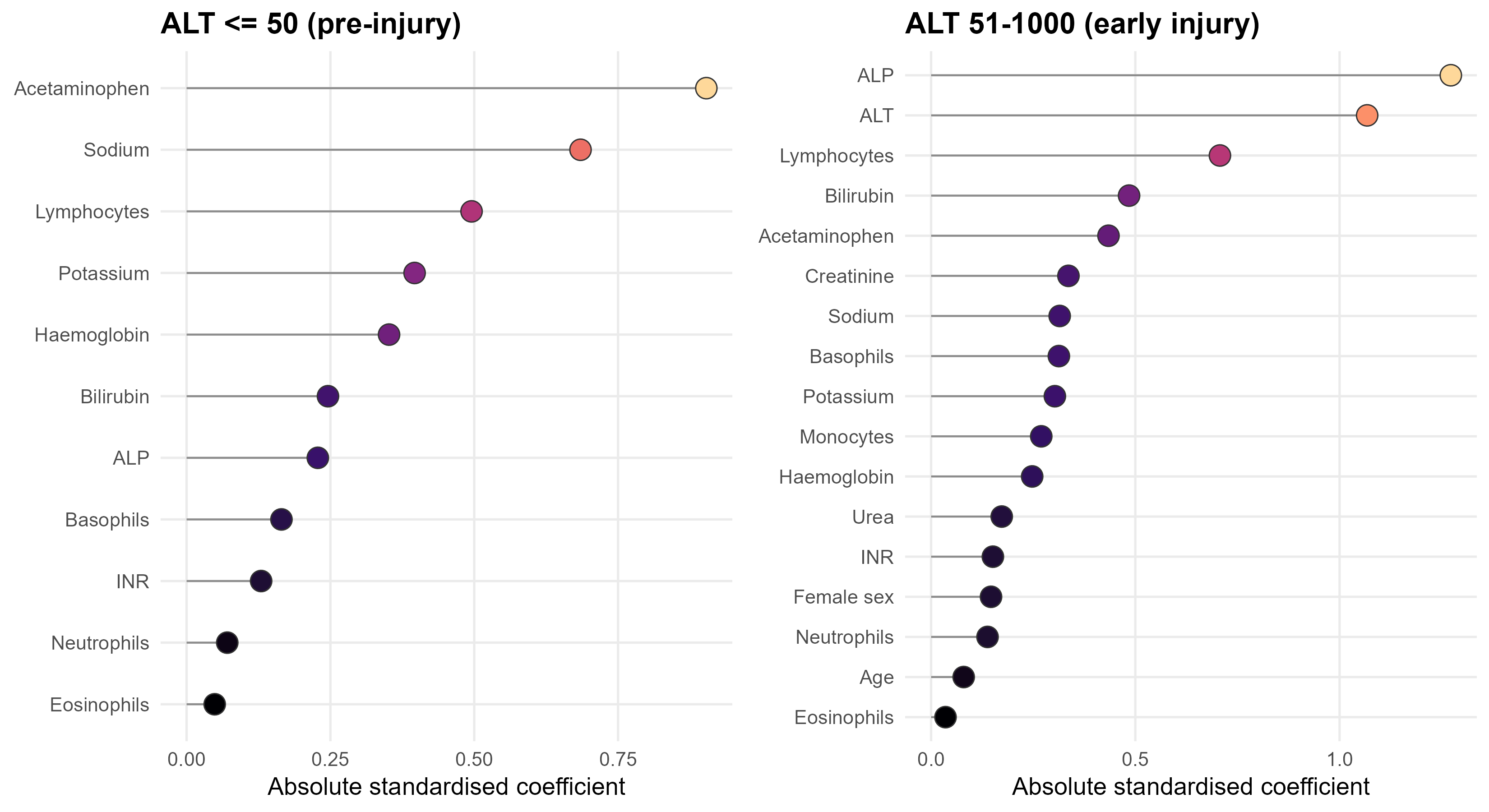

Figure E4b. Predictor importance (absolute standardised coefficient) in the full model, by stratum; this defines the order in which predictors entered the nested models. Pre-injury stratum on the left.

### E5. Discrimination and per-stratum calibration

In the held-out test set the parsimonious model discriminated hepatotoxicity with an AUROC of 0.93 (95% CI 0.89 to 0.97) against 0.82 (95% CI 0.72 to 0.91) for the ALTxAPAP rule, a difference of 0.11 (95% CI 0.01 to 0.22; paired stratified bootstrap of 2,000 replicates, p = 0.03; the bootstrap is used in preference to DeLong's asymptotic test because the test set contains only 29 events; pROC). The full 17-predictor model reached an AUROC of 0.94 (95% CI 0.90 to 0.98) and the selected four-per-stratum model 0.93, so the seven-test model retained the discrimination of the full model while using far fewer inputs.

| Model | Predictors | AUROC (95% CI) |
| --- | --- | --- |
| Edinburgh Risk Score | 7 distinct (4 per stratum) | 0.93 (0.89 to 0.97) |
| ALTxAPAP rule | 2 | 0.82 (0.72 to 0.91) |
| Difference (model minus ALTxAPAP) | - | 0.11 (0.01 to 0.22) |
| Seventeen-biomarker model | 17 | 0.94 (0.90 to 0.98) |

Calibration is reported within each stratum rather than as a single pooled slope, because the model is fitted and calibrated separately in each stratum. In the early-injury stratum (ALT 51 to 1,000; n 156, 22 test events) the calibration slope was 1.45 (95% CI 0.87 to 2.81), calibration-in-the-large was +0.29 and the Brier score was 0.092. The pre-injury stratum (ALT at or below 50; n 1019, 7 test events) had a point slope of 0.98 with calibration-in-the-large -0.22 and a Brier score of 0.007; with only 7 events its interval is very wide, so the slope is shown below for completeness but should be read with caution rather than as evidence of good calibration. The per-stratum training event counts were 24 in the pre-injury stratum (ALT at or below 50) and 66 in the early-injury stratum (ALT 51 to 1,000).

| Stratum | N | Test events | Calibration slope (95% CI) | CITL | Brier |
| --- | --- | --- | --- | --- | --- |
| Early injury (ALT 51 to 1,000) | 156 | 22 | 1.45 (0.87 to 2.81) | +0.29 | 0.092 |
| Pre-injury (ALT at or below 50) | 1019 | 7 | 0.98 (wide; 7 events, read with caution) | -0.22 | 0.007 |

Because calibration carries real uncertainty at this event count, the operating point in section E8 is chosen on ranking (sensitivity matched to current practice) rather than on a calibrated risk value, and the decision curve in section E7 is reported as supporting evidence only.

### E6. Risk refinement: integrated discrimination improvement and categorical reclassification

Risk refinement was assessed by the integrated discrimination improvement, a threshold-free measure that uses no operating point and imposes no risk cut on either rule, so it summarises how the model separates predicted risk between events and non-events relative to the comparator across the whole probability scale rather than at a single point. The comparator risk was the ALTxAPAP risk from a logistic model fitted on the training set on the log scale, so the comparison is leakage-free. The integrated discrimination improvement was 0.13. It is reference-dependent, being defined relative to the ALTxAPAP comparator, so we report it as supporting rather than primary.

Reclassification itself is reported only as the bounded categorical comparison at the matched-sensitivity operating point, cross-tabulating the model (high or low at the matched point) against current ALTxAPAP > 1,500 practice in the 1,175-patient test set with 29 events. Because both rules are pinned to the same sensitivity, they identify the same number of patients who progressed (26 of 29), and the small event-level discordance (three patients each way) cancels, so the event component is zero and the reclassification gain falls on the non-events: the model returns to low risk 303 of the non-events that ALTxAPAP flags while wrongly flagging only 75 the other way. The categorical net reclassification index therefore totals 0.20, with an event component of 0 and a non-event component of +0.20. The full two-by-two cross-tabulation underlying this comparison is given with the operating characteristics in section E8.

| Categorical reclassification (matched-sensitivity point vs ALTxAPAP > 1,500) | Event component | Non-event component | Total |
| --- | --- | --- | --- |
| Categorical NRI | 0 | +0.20 | 0.20 |
| IDI (threshold-free) | - | - | 0.13 |

### E7. Decision-curve analysis

Across the range of credible decision thresholds, the net benefit of the parsimonious model exceeded that of the ALTxAPAP rule and of the treat-all and treat-none strategies. This is shown in panel B of main-text Figure 2. Net benefit assumes good calibration, so consistent with section E5 we treat the decision curve as supporting evidence for the model's clinical utility rather than as the headline result; the matched-sensitivity dominance in section E8, which depends only on ranking, carries the primary argument.

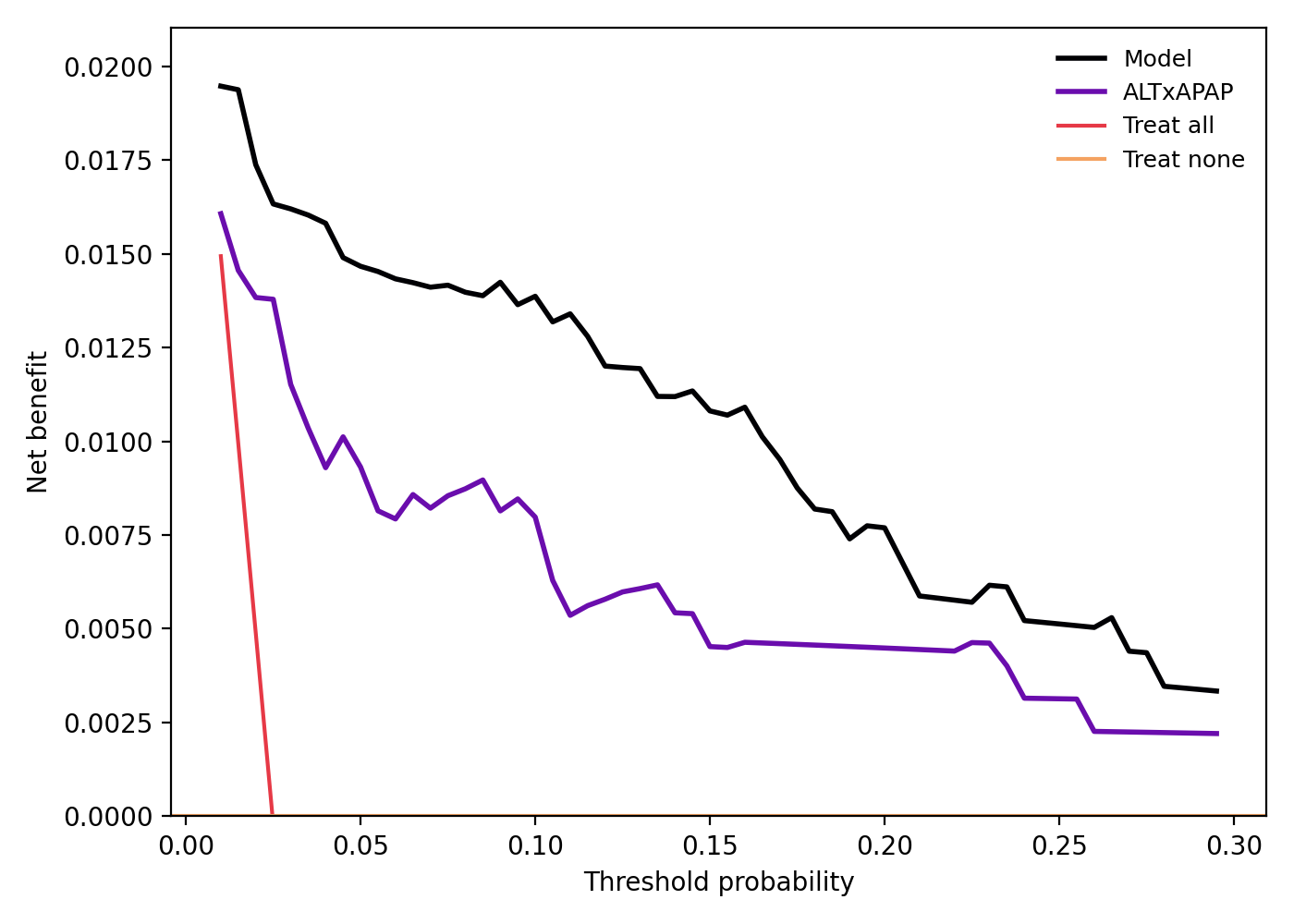

Figure E7. Decision-curve analysis: net benefit of the model versus the ALTxAPAP comparator across threshold probabilities.

### E8. Operating characteristics: matched-sensitivity head-to-head and operating-point menu

Against each conventional ALTxAPAP threshold, the model was set to the comparator's sensitivity at that cut-off and compared at that matched sensitivity, improving specificity, raising the positive likelihood ratio and cutting false positives in the 1,175-patient test set. At the sensitivity of the ALTxAPAP product above 1,500 (89.7%), the ALTxAPAP rule gave specificity 62.6%, positive likelihood ratio 2.39, negative likelihood ratio 0.17, with 429 false positives and 455 patients flagged; the model at matched sensitivity gave specificity 82.5%, positive likelihood ratio 5.11, negative likelihood ratio 0.13, with 201 false positives and 227 patients flagged, a 53% reduction in false positives (201 vs 429). At the ALTxAPAP product above 3,000 (sensitivity 65.5%), the ALTxAPAP rule gave specificity 81.8%, positive likelihood ratio 3.61 and negative likelihood ratio 0.42, against model specificity 97.5%, positive likelihood ratio 25.9 and negative likelihood ratio 0.35. At the ALTxAPAP product above 10,000 (sensitivity 41.4%), the ALTxAPAP rule gave specificity 98.0%, positive likelihood ratio 20.6 and negative likelihood ratio 0.60, against model specificity 99.0%, positive likelihood ratio 43.1 and negative likelihood ratio 0.59.

| Comparison (test set, n=1,175) | Sensitivity | Specificity | LR+ | LR- | False positives |
| --- | --- | --- | --- | --- | --- |
| ALTxAPAP > 1,500 | 89.7% | 62.6% | 2.39 | 0.17 | 429 |
| Model at matched sensitivity | 89.7% | 82.5% | 5.11 | 0.13 | 201 |
| ALTxAPAP > 3,000 | 65.5% | 81.8% | 3.61 | 0.42 | - |
| Model at matched sensitivity | 65.5% | 97.5% | 25.9 | 0.35 | - |
| ALTxAPAP > 10,000 | 41.4% | 98.0% | 20.6 | 0.60 | - |
| Model at matched sensitivity | 41.4% | 99.0% | 43.1 | 0.59 | - |

Because the model can be read at any threshold, the table below offers an operating-point menu spanning the clinically useful range of sensitivities. Each row gives the calibrated probability cut, the resulting sensitivity and specificity with 95% confidence intervals, the positive and negative likelihood ratios, and the number of the 1,175 test patients flagged. The row matched to current ALTxAPAP > 1,500 practice identifies about 90% of patients who progress while more than halving the false positives of the existing rule.

| Operating point / basis | Threshold | Sensitivity (95% CI) | Specificity (95% CI) | LR+ | LR- | Flagged |
| --- | --- | --- | --- | --- | --- | --- |
| 75% sensitivity | 9.4% | 0.76 (0.58 to 0.88) | 0.96 (0.94 to 0.97) | 18.1 | 0.25 | 70 |
| 80% sensitivity | 2.1% | 0.83 (0.65 to 0.92) | 0.84 (0.82 to 0.86) | 5.30 | 0.20 | 203 |
| Matched to ALTxAPAP > 1,500 | 1.7% | 0.90 (0.74 to 0.96) | 0.82 (0.80 to 0.85) | 5.11 | 0.13 | 227 |
| 90% sensitivity | 0.9% | 0.93 (0.78 to 0.98) | 0.67 (0.64 to 0.69) | 2.79 | 0.10 | 409 |
| 95% sensitivity | 0.8% | 0.97 (0.83 to 0.99) | 0.63 (0.61 to 0.66) | 2.64 | 0.05 | 447 |

At the matched-sensitivity operating point the model can also be cross-tabulated against current ALTxAPAP > 1,500 practice. Because the two rules are compared at the same sensitivity, they flag the same number of patients who progressed but not the identical patients. Among the 29 events, 23 were flagged high by both, 3 by the model only, 3 by ALTxAPAP only and 0 by neither, so each rule caught 26 of 29 but they disagreed on six. Among the non-events, 126 were flagged high by both, 75 by the model only, 303 by ALTxAPAP only and 642 by neither, so the model returned to low risk 303 of the non-events that ALTxAPAP flags while moving only 75 the other way. This is the bounded categorical reclassification summarised in section E6 (categorical NRI 0.20, event component 0, non-event component +0.20).

| Reclassification 2x2 (model at matched sensitivity vs ALTxAPAP > 1,500) | Both high | Model only | ALTxAPAP only | Both low |
| --- | --- | --- | --- | --- |
| Events (n=29) | 23 | 3 | 3 | 0 |
| Non-events | 126 | 75 | 303 | 642 |

### E9. Duplicate-inclusive sensitivity analyses

The primary analysis retained one admission per patient so the model could not learn an individual patient's recurring biochemistry. To check that this decision was sound, we re-analysed the full encounter-level cohort (6,378 admissions from 4,705 patients, 121 events, 1.90%) using a mixed-effects logistic model with a patient-level random intercept. The patient random-intercept variance was 2.34 on the logit scale, which is substantial, indicating that repeat admissions are correlated and supporting the decision to retain one admission per patient. The mixed model was fit on a single completed imputation, because stacking the 50 imputations collapses the random-effect variance. A pooled logistic model with patient-clustered (cluster-robust) standard errors gave coefficient directions consistent with the primary model, with acetaminophen, ALT, lymphocyte count, alkaline phosphatase, sodium and potassium all retaining their expected signs. Predictor directions are consistent under the random intercept and under patient-clustered standard errors.

| Quantity | Value |
| --- | --- |
| Encounter-level admissions | 6,378 |
| Distinct patients | 4,705 |
| Events | 121 (1.90%) |
| Patient random-intercept variance (logit scale) | 2.34 (substantial) |

### E10. Missing data and performance by completeness

#### Missingness mechanism and the missing-not-at-random concern

Laboratory panels are ordered together, so an unordered full blood count removes all of its components at once, and similarly for urea-and-electrolytes and liver function tests. Because the decision to order a panel is itself clinical, the missingness is plausibly missing not at random (MNAR): whether a value is observed may depend on the clinician's unrecorded judgement of how unwell the patient is, which is correlated with the value that would have been recorded. Rather than impose a particular MNAR departure on the imputed values, we judged robustness empirically, by reporting performance separately in patients with complete and with imputed data (below).

We mitigated this in two ways. Multiple imputation by chained equations was specified with a rich predictor set and the outcome included in the imputation model, which recovers block missingness from the jointly observed panels and makes the missing-at-random assumption more plausible conditional on the observed data: where one component of a panel is missing it is rarely missing in isolation, and where it is, the co-ordered values and the outcome carry most of the information needed to impute it. Per-variable missingness is reported in full below so the reader can judge where imputation does the most work. INR had the highest missingness at 20.1% and eosinophil count the next highest at 9.5%; the predictors used by the primary model were each missing in no more than 4.1% of admissions (acetaminophen 3.3%, lymphocyte count 3.1%, potassium 4.1%, sodium 0.7%, alkaline phosphatase 0.1%, bilirubin 0.0%, ALT 0.0%).

| Variable | Missing (n) | Missing (%) |
| --- | --- | --- |
| INR | 946 | 20.1 |
| Eosinophil count | 447 | 9.5 |
| Basophil count | 195 | 4.1 |
| Potassium | 191 | 4.1 |
| Acetaminophen | 157 | 3.3 |
| Haemoglobin | 151 | 3.2 |
| Lymphocyte count | 145 | 3.1 |
| Monocyte count | 146 | 3.1 |
| Neutrophil count | 145 | 3.1 |
| Sodium | 31 | 0.7 |
| Urea | 33 | 0.7 |
| Creatinine | 29 | 0.6 |
| Alkaline phosphatase | 5 | 0.1 |
| ALT | 0 | 0.0 |
| Bilirubin | 0 | 0.0 |
| Sex | 2 | 0.0 |
| Age | 0 | 0.0 |

#### Missingness by presentation-ALT stratum

Because laboratory panels are ordered differently in pre-injury and early-injury presentations, missingness is also reported within each stratum (pre-injury n 4,080; early injury n 625). Every model predictor was missing in no more than about 4% of admissions in either stratum; INR and eosinophil count, neither of which is a model predictor, carried the most missingness.

| Variable | Pre-injury (ALT<=50) missing % | Early injury (51-1000) missing % |
| --- | --- | --- |
| INR | 20.6 | 16.6 |
| Eosinophil count | 8.4 | 16.6 |
| Potassium | 4.2 | 2.9 |
| Lymphocyte count | 3.0 | 3.8 |
| Acetaminophen | 3.3 | 3.4 |
| Sodium | 0.7 | 0.6 |
| Alkaline phosphatase | 0.1 | 0.0 |
| ALT | 0.0 | 0.0 |
| Bilirubin | 0.0 | 0.0 |

#### Performance stratified by data completeness

We next compared model performance in patients whose required data were complete against those who would need imputation to be scored. Completeness is judged on the four predictors that drive each patient’s own stratum model, since those are the only values the deployed score requires; a patient missing an unrelated laboratory value is not counted as incomplete. Of the 1,175 test patients, 1,067 (91%) had all four required predictors observed, so their score was computed with no imputation; in this group the AUROC was 0.94, with sensitivity 0.93 and specificity 0.82 at the matched-sensitivity operating point. This complete-case estimate, which involves no imputation at all, is the deployment-relevant figure and is consistent with the full-cohort result. Only 108 patients (2 events) were missing a required predictor; with so few events no meaningful performance estimate is possible in this subgroup, and because any score there depends on imputation we do not interpret it and cannot exclude systematically biased predictions for such patients. The observed event rate did not differ materially by completeness (2.5% versus 1.9%), so we do not read missingness as prognostic.

| Group | N | Prevalence | Sensitivity | Specificity | AUROC |
| --- | --- | --- | --- | --- | --- |
| Complete (all four required predictors) | 1,067 | 2.5% (27 events) | 0.93 | 0.82 | 0.94 |
| Missing ≥1 required predictor | 108 | 1.9% (2 events) | 0.50 | 0.92 | 0.87 |

### E11. Acetylcysteine protocol comparison

The cohort spans two acetylcysteine regimens, the 21-hour regimen and the SNAP regimen. The model discriminated hepatotoxicity in patients treated under each regimen: AUROC 0.94 under the 21-hour regimen (n = 660, 15 events) and 0.92 under SNAP (n = 515, 14 events). For the purposes of this study the relevance is that the relationship between the admission bloods and the outcome holds under both regimens, so the model can be developed on patients given one regimen and applied to patients given the other. We did not set out to compare the regimens themselves and draw no conclusion here about their relative effectiveness. The risk model applies regardless of which acetylcysteine regimen is used.

| Regimen | N | Events | AUROC |
| --- | --- | --- | --- |
| 21-hour | 660 | 15 | 0.94 |
| SNAP | 515 | 14 | 0.92 |

### E12. Reproducibility

Analysis code is provided as R Markdown modules with a single global seed. A data-free public copy is included; NHS Lothian patient data cannot be shared publicly (Caldicott approval ref. 24167). A TRIPOD-AI checklist accompanies the submission, and the model-size selection rationale is described in section E4.

The analysis code is archived at Zenodo (doi: 10.5281/zenodo.21358187).
