## Supplementary material for "Prediction of acetaminophen-induced hepatotoxicity in acetylcysteine-treated patients using routine admission biomarkers": Reporting checklist

TRIPOD+AI reporting checklist

Prediction of acetaminophen-induced hepatotoxicity in acetylcysteine-treated patients using routine admission biomarkers (Annals of Emergency Medicine, MS 2026-719). Checklist per Collins GS, et al. TRIPOD+AI statement. BMJ 2024;385:e078378. D=development, E=evaluation; this is a development with internal validation. Rows shaded in peach require a declaration to be added to the manuscript title page.

### Main checklist (27 items)

| **Item** | **Checklist item (abridged; see TRIPOD+AI, BMJ 2024;385:e078378)** | **Reported (location in this submission)** |
| --- | --- | --- |
| **Title and Abstract** | | |
| 1 | Identify the study as developing/evaluating a multivariable prediction model, the target population, and the outcome. | Title |
| 2 | Structured abstract (see TRIPOD+AI for Abstracts, below). | Abstract |
| **Introduction** | | |
| 3a | Healthcare context and rationale, including references to existing models. | Introduction — Background & Importance (ALT×APAP) |
| 3b | Target population and intended purpose in the care pathway, including intended users. | Introduction — Importance & Goals; Discussion |
| 3c | Any known health inequalities between sociodemographic groups. | Not formally assessed; cohort case-mix (young, female) noted in Limitations |
| 4 | Study objectives (development, validation, or both). | Introduction — Goals of this investigation (development + internal validation) |
| **Methods** | | |
| 5a | Sources of data (development/evaluation), rationale, representativeness. | Methods — Design & setting; Selection of participants; Appendix E1 |
| 5b | Dates of participant accrual and, if applicable, end of follow-up. | Methods — Design & setting (March 2008–October 2024) |
| 6a | Study setting; number and location of centres. | Methods — Design & setting (three NHS Lothian hospitals) |
| 6b | Eligibility criteria for participants. | Methods — Selection of participants; Appendix E1 |
| 6c | Treatments received and how handled during development/evaluation. | Methods; Table 1 (acetylcysteine regimen); Appendix E11 |
| 7 | Data pre-processing and quality checks. | Methods — Measurements; Appendix E2 (availability filter, exclusions, INR relabel) |
| 8a | Outcome definition, time horizon, rationale, consistency of assessment. | Methods — Outcomes (peak ALT >1,000 U/L after first ALT); Appendix E1 (endpoint validation) |
| 8b | Qualifications of outcome assessors (if subjective). | Not applicable — objective laboratory outcome (peak ALT) |
| 8c | Actions to blind outcome assessment. | Not applicable — objective laboratory outcome, retrospective |
| 9a | Choice of initial predictors and any pre-selection. | Methods — Measurements & Analysis; Appendix E2/E4; Figure 3 (model-size) |
| 9b | Definition of all predictors, how/when measured, blinding. | Methods — Measurements (±2 h of first ALT); Table 1 |
| 9c | Qualifications of predictor assessors (if subjective). | Not applicable — objective laboratory predictors |
| 10 | How study size was arrived at and justification. | Full available cohort; 75/25 patient-level split; Riley et al. minimum-sample-size criteria checked (Appendix E4). No a priori power calculation |
| 11 | How missing data were handled. | Methods — Measurements (MICE, 50 imputations, re-fitted within CV folds); Appendices E2, E11; Limitations |
| 12a | How data were used/partitioned in the analysis. | Methods — Analysis (patient-level 75/25 split; test set held out from all development) |
| 12b | How predictors were handled (functional form, rescaling, standardisation). | Methods/Appendix E2 (centre & scale; elastic-net); Appendix E3 (raw-value equation) |
| 12c | Model type, rationale, all building steps, hyperparameter tuning, internal validation. | Methods — Analysis; Appendix E2 (elastic-net glmnet/caret; α∈[0.1,0.9], λ grid; 10-fold patient-grouped CV); Appendix E4 (size selection) |
| 12d | Heterogeneity in estimates/performance across clusters. | Appendix E10 (patient random intercept & cluster-robust SEs); E12 (protocol subgroups) |
| 12e | Measures/plots used to evaluate performance. | Methods — Analysis; Figures 2–3; Appendices E5–E8 (AUROC, calibration, DCA, IDI, reclassification) |
| 12f | Any model updating/recalibration. | Platt recalibration per stratum — Methods; Appendix E2/E3 |
| 12g | How model predictions were calculated for evaluation. | Table 2 (equation); Appendix E3; Excel calculator |
| 13 | Class-imbalance methods, and any subsequent recalibration. | Methods — Analysis (class-balanced weighting); Platt recalibration; Appendix E2 |
| 14 | Approaches used to address model fairness. | Paediatric and acetylcysteine-protocol subgroups (Results; Appendix E12). Formal sociodemographic fairness analysis not performed — single-region cohort |
| 15 | Model output and any classification thresholds. | Results — Operating characteristics; Table 3 (operating-point menu by target sensitivity); Appendix E8 |
| 16 | Differences between development and evaluation data. | Not applicable — internal validation; training and held-out test set drawn from the same cohort |
| 17 | Ethics approval and consent or waiver. | Statements |
| **Open science** | | |
| 18a | Source of funding and role of funders. | Statements |
| 18b | Conflicts of interest / financial disclosures. | Statements |
| 18c | Where the protocol can be accessed, or that none was prepared. | No protocol was prepared – the study was registered prospectively with the study sponsor for ethical approval. |
| 18d | Study registration (register name and number) or state not registered. | No protocol was prepared – the study was registered prospectively with the study sponsor for ethical approval. |
| 18e | Availability of the study data. | Methods; Appendix E13 — NHS Lothian data Caldicott-restricted, cannot be shared; data-free public code release |
| 18f | Availability of the analytical code. | Methods — Analysis (Zenodo, DOI to be minted on acceptance); Appendix E13 (PUBLIC_release modules) |
| **Patient and public involvement** | | |
| 19 | Any patient/public involvement, or state none. | None |
| **Results** | | |
| 20a | Flow of participants (numbers with/without outcome; follow-up). | Figure 1; Appendix E1 |
| 20b | Characteristics overall and by source/setting; key dates, predictors, events, missing data. | Table 1 (by outcome and by training/test set) |
| 20c | Comparison of predictor/outcome distribution (evaluation vs development). | Table 1 (training vs test distributions) |
| 21 | Number of participants and events in each analysis. | Table 1; Figure 1; Results; per-stratum training events (Appendix E4) |
| 22 | Full model specification to allow prediction in new individuals. | Table 2 (equation, coefficients, Platt intercept/slope); Appendix E3; calculator; Zenodo code |
| 23a | Performance estimates with confidence intervals, incl. key subgroups. | Results (AUROC & operating characteristics with 95% CIs); Table 3; paediatric/protocol subgroups; Appendices E5–E8 |
| 23b | Heterogeneity in performance across clusters, if examined. | Appendix E10; E12 |
| 24 | Results of any model updating. | Not applicable — no external model updating |
| **Discussion** | | |
| 25 | Overall interpretation, including fairness, vs objectives and prior work. | Discussion |
| 26 | Limitations (representativeness, sample size, overfitting, missing data) and effects. | Limitations |
| 27a | How poor-quality/unavailable input data should be handled at implementation. | Limitations & Appendix E11 — score requires the four stratum predictors; complete-case is the deployment-relevant estimate |
| 27b | Whether users interact with input/model and expertise required. | Discussion — Implementation; Table 2; Excel calculator + companion note |
| 27c | Next steps for future research (applicability, generalisability). | Discussion (external & prospective validation; cost-effectiveness) |

### TRIPOD+AI for Abstracts (13 items)

| **Item** | **TRIPOD+AI for Abstracts item** | **Reported (Abstract)** |
| --- | --- | --- |
| 1 | Identify study as developing/evaluating a prediction model, population, outcome. | Title |
| 2 | Brief healthcare context and rationale. | Abstract — Study objective |
| 3 | Objectives (development, evaluation, or both). | Abstract — Study objective |
| 4 | Sources of data. | Abstract — Methods |
| 5 | Eligibility criteria and setting. | Abstract — Methods |
| 6 | Outcome, including time horizon. | Abstract — Methods (peak ALT >1,000 U/L after first ALT) |
| 7 | Model type, model-building summary, internal validation. | Abstract — Methods (elastic-net, ALT-stratified; 25% held-out test) |
| 8 | Measures used to assess performance. | Abstract — Methods/Results (AUROC, specificity, likelihood ratio) |
| 9 | Number of participants and events. | Abstract — Results (4,705 admissions; test n=1,175, 29 events) |
| 10 | Predictors in the final model. | Abstract — Results (seven routine tests, four per stratum) |
| 11 | Performance estimates with confidence intervals. | Abstract — Results (AUROC 0.93 [0.89–0.97] vs 0.82 [0.72–0.91]) |
| 12 | Overall interpretation. | Abstract — Conclusion |
| 13 | Registration number and registry/repository. | Not registered; analytical code at Zenodo (DOI on acceptance) |
